# Exploring Unique Device Identifier Implementation and Use for Real-World Evidence: A Mixed-Methods Study with NESTcc Health System Network Collaborators

**DOI:** 10.1101/2022.09.21.22279992

**Authors:** Sanket S. Dhruva, Jennifer L. Ridgeway, Joseph S. Ross, Joseph P. Drozda, Natalia A. Wilson

## Abstract

**Objectives:** To examine the current state of Unique Device Identifier (UDI) implementation, including barriers and facilitators, among eight health systems participating in a research network committed to real-world evidence (RWE) generation for medical devices.

**Design:** Mixed methods, including a structured survey and semi-structured interviews.

**Setting:** Eight health systems participating in the National Evaluation System for health Technology research network within the United States.

**Participants:** Individuals identified as being involved in or knowledgeable about UDI implementation or medical device identification from supply chain, information technology, and high-volume procedural area(s) in their health system.

**Main Outcomes Measures:** Interview topics were related to UDI implementation, including barriers and facilitators; UDI use; benefits of UDI adoption; and vision for UDI implementation. Data were analyzed using directed content analysis, drawing on prior conceptual models of UDI implementation and the Exploration, Preparation, Implementation, Sustainment (EPIS) framework. A brief survey of health system characteristics and scope of UDI implementation was also conducted.

**Results:** Thirty-five individuals completed interviews. Three of eight health systems reported having implemented UDI. Themes identified about barriers and facilitators to UDI implementation included knowledge of the UDI and its benefits among decision makers; organizational systems, culture, and networks that support technology and workflow changes; and external factors such as policy mandates and technology. A final theme focused on the availability of UDIs for RWE; lack of availability significantly hindered RWE studies on medical devices.

**Conclusions:** UDI adoption within health systems requires knowledge of and impetus to achieve operational and clinical benefits. These are necessary to support UDI availability for medical device safety and effectiveness studies and RWE generation.

**Key Messages:** *What is already known on this topic:*

- Recent legislation and policy have placed increased emphasis on tracking medical device safety and effectiveness using real-world evidence (RWE).
- The unique device identifier (UDI), available on labels of most moderate- and high-risk medical devices, ensures accurate and reliable medical device identification and tracking.

*What this study adds:*

- Even among health systems committed to RWE generation, UDIs are often not available within their data sources to enable effective medical device identification for RWE studies.
- Knowledge by health system leaders about UDI benefits and buy-in about its operational and clinical benefits are necessary for UDI implementation and availability for RWE generation.

*How this study might affect research, practice or policy:*

- Results demonstrate the need to increase awareness and provide guidance about the value of UDI use to health system leadership.
- Policy mandates are necessary to drive greater UDI adoption in health systems and support real-world evidence generation.

## INTRODUCTION

In recent years, both legislation and U.S. Food and Drug Administration (FDA) policy have placed an increasing emphasis on the use of real-world evidence (RWE) to support evaluations of medical device safety and effectiveness, including for regulatory decision-making.^1 2^ RWE is clinical evidence synthesized from data captured from sources other than traditional clinical research. Real-world data (RWD) sources that can contribute to RWE include, but are not limited to, electronic health records (EHRs), insurance claims, and registries.^3^ RWD have several advantages compared with traditional research data sources: they are ubiquitously available, less expensive, and available for more diverse patient populations than usually represented in clinical trials.^4 5^

To drive the quality and efficiency in use of RWD for medical device evaluation, FDA established the National Evaluation System for health Technology Coordinating Center (NESTcc).^6^ NESTcc leverages its research network collaborators to generate high-quality RWE to inform medical device decision-making by regulators, payors, and clinicians, for the ultimate benefit of patient safety and quality of care.^7-9^ A fundamental necessity to support this work is that medical devices must be accurately identified and linked to the patients in whose care they are used.

Key legislative steps in this direction were the 2007 FDA Amendments Act and then the 2012 FDA Safety and Innovation Act, which mandated FDA to publish regulations to establish a system where Unique Device Identifiers (UDIs) would be available for medical device identification.^10 11^ A key impetus was significant medical device-related adverse events.^7 12^ Subsequently, FDA published the 2013 UDI System Rule, which mandated a distinct code on the label and packaging of medical devices marketed in the United States.^13 14^ The UDI contains both a device identifier, which includes the manufacturer’s name and the model of the specific device; and a production identifier, which includes, as available, the lot and serial number, manufacturing date, and expiration date. Manufacturers have complied with this mandate and most moderate and high-risk devices are labeled with a UDI.

However, for the UDI to be leveraged for RWE generation, it must be integrated into structured fields within electronic health information technology (IT) systems – primarily EHRs – when a medical device is used in a patient’s care. UDI integration is a critical step in ensuring these data are available for transmission to research databases and in claims forms, thus available for downstream use. Limited availability of UDIs in RWD has been cited as a significant data challenge in generating RWE.^15-17^ Although the Office of the National Coordinator for Health Information Technology requires inclusion of the UDI for implantable medical devices for EHR certification, with an appropriate field, there is no mandate for hospitals to populate this field.^18^ While some health systems have delineated the value of this integration clinically and operationally for their organization and have implemented UDI capture into health IT systems at the point-of-care,^19 20^ unfortunately most have not^21^ in the setting of organizational, workflow, and information technology challenges.^22^

Understanding how health systems perceive the value of UDI and choose to implement it as a structured data element within an EHR is an essential next step to support strategies to increase the availability of medical device data for RWD-based studies. Prior research on health systems that had implemented UDI for implantable devices informed a roadmap for health system implementation as well as barriers, strategies, and next steps.^21 23^ This project sought to build on this work and characterize the current state of UDI implementation and use in a focused group of health systems, those affiliated with NESTcc. Ultimately, these findings are intended to inform a playbook for UDI implementation in NESTcc health system network collaborators and for broader use by health systems, to support advancement of RWE on medical devices.

## METHODS

### Setting and Study Oversight

This project was supported by NESTcc, an organization with a network of research collaborators focused on generating RWE for medical device testing, approval, and monitoring.^8^ The study team included investigators with expertise in UDI implementation, medical device evaluation, use of RWD, and qualitative methods. The study team met with NESTcc leadership monthly. This project received institutional review board approval.

### Interviewee Recruitment and Pre-Interview Survey

All U.S.-based health systems and clinical research networks (CRNs) affiliated with NESTcc as of March 2021 were eligible for inclusion. Among CRNs, a single health system was included. For each identified health system, the NESTcc Senior Vice President sent an initial email to the NESTcc network collaborator lead, introducing the study and requesting participation. The study team sent a follow-up email, including a brief pre-interview survey (**Supplementary File 1**) querying basic information about the health system, scope of UDI implementation, and names of individuals focused on UDI implementation or medical device identification from supply chain, IT, and high-volume procedural area(s), as well as any UDI champion. Surveys were returned by email and responses were entered into a REDCap database for analysis.

Individuals recommended for interviews received an invitation by email. Those who agreed to participate were scheduled for a one-hour semi-structured interview. The aim was to recruit three individuals from a variety of UDI-related roles (IT, supply chain, clinical leadership) in each health system. A minimum of three contact attempts were made per individual. Chain-referral sampling subsequently was used to identify additional individuals appropriate to interview.

### Semi-Structured Interview Guide and Interviews

The study team developed a semi-structured interview guide, which was informed by the literature on UDI implementation and use;^20 21 24-26^ constructs from implementation science for personal, organizational, and external factors related to implementation;^27 28^ and study team member expertise. Topics in the guide included UDI implementation, barriers and facilitators, use, and benefits of adoption. If UDI was unavailable, questions focused on understanding why, as well as the desire/vision to implement UDI. The guide underwent public comment through NESTcc. Feedback was reviewed by the study team and informed the final guide. (**Supplementary File 2**).

### Data Collection

Interviews were conducted via videoconference between May and October 2021 by at least two members of the study team experienced in qualitative interviewing techniques. All participants provided oral consent prior to the start of the interview. Interviews were recorded with permission, transcribed verbatim, and de-identified. Interviewers completed field notes for each interview. Impressions were presented to the study team for feedback at bimonthly study meetings. All data were stored on a secure server and available only to members of the study team.

### Data Analysis

Survey data were aggregated and summarized in a spreadsheet. Health system demographic variables were supplemented using American Hospital Directory data.^29^ Qualitative analysis employed methods of directed content analysis.^30^ A coding framework derived from existing conceptual models of UDI implementation and the Exploration, Preparation, Implementation, Sustainment (EPIS) implementation framework^21 28^ was developed, followed by review of the first five transcripts to identify any additional constructs or topics. Two members of the study team then independently coded the first 25% of transcripts. Resulting coding was compared for concordance, which was 87.6%. Differences were reviewed to ensure common understanding of the coding framework, and codebook definitions and transcript coding were updated. Given high concordance, the study team members independently coded the remaining transcripts. Coded transcripts were entered into qualitative data analysis software (NVivo, QSR International) to facilitate queries for analysis. Findings were organized into themes and subthemes that represent the personal, organizational, and external factors that support or inhibit UDI implementation and experiences with UDI availability for RWE studies. Members of the larger study team reviewed findings and coded excerpts as a way of reducing bias in the analysis process.

## RESULTS

### Participation

Eight of 12 eligible health systems participated by completing at least one interview.

This included six individual NESTcc health system network collaborators and two health system members of a NESTcc CRN network collaborator.

### Pre-Interview Surveys

Demographics of the eight participating health systems are shown in **Table 1**. All were not-for-profit. The number of hospitals per health system ranged from 1 to 43, with a median of 1,795 licensed beds per health system.

**Table 1.**
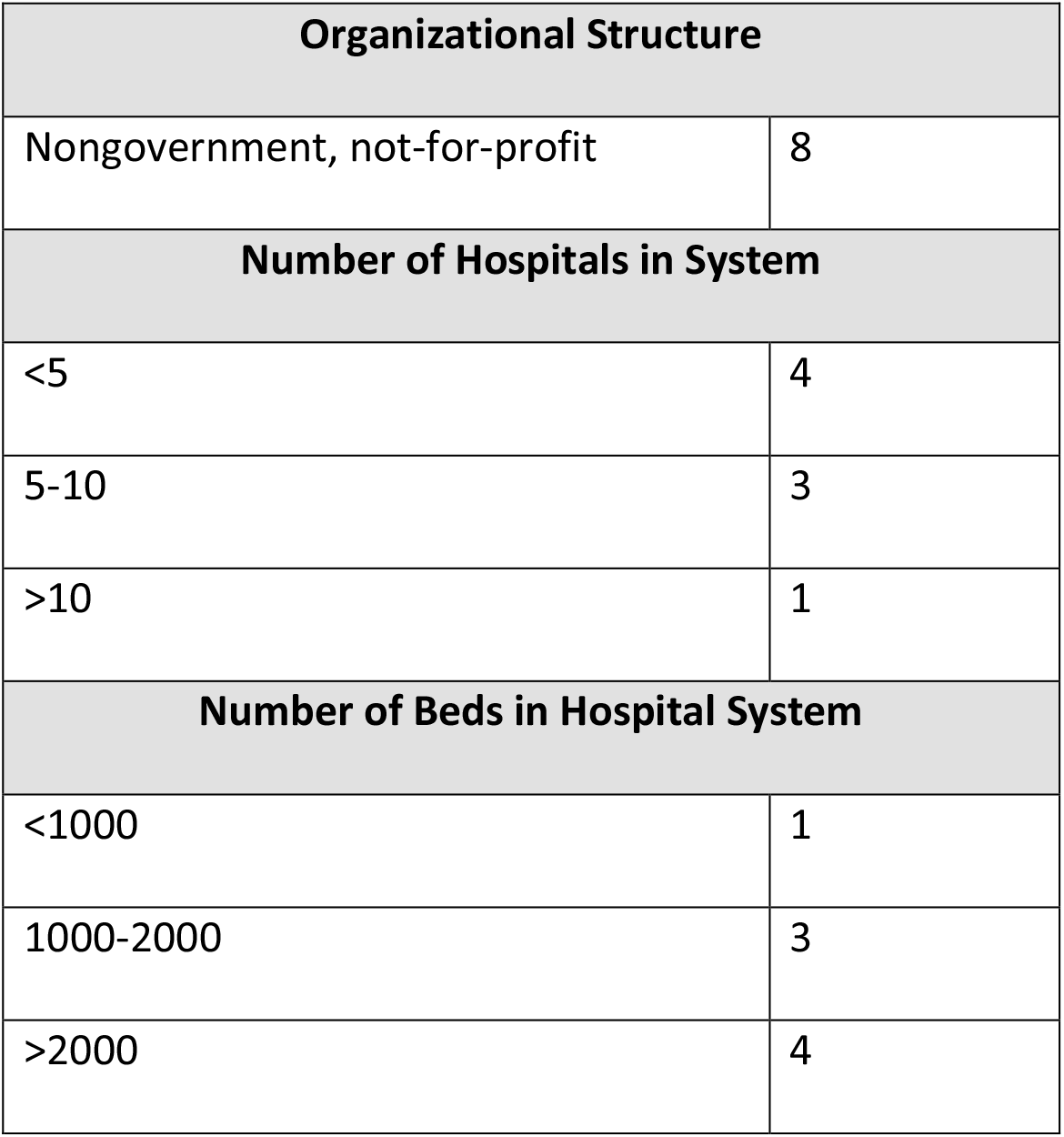
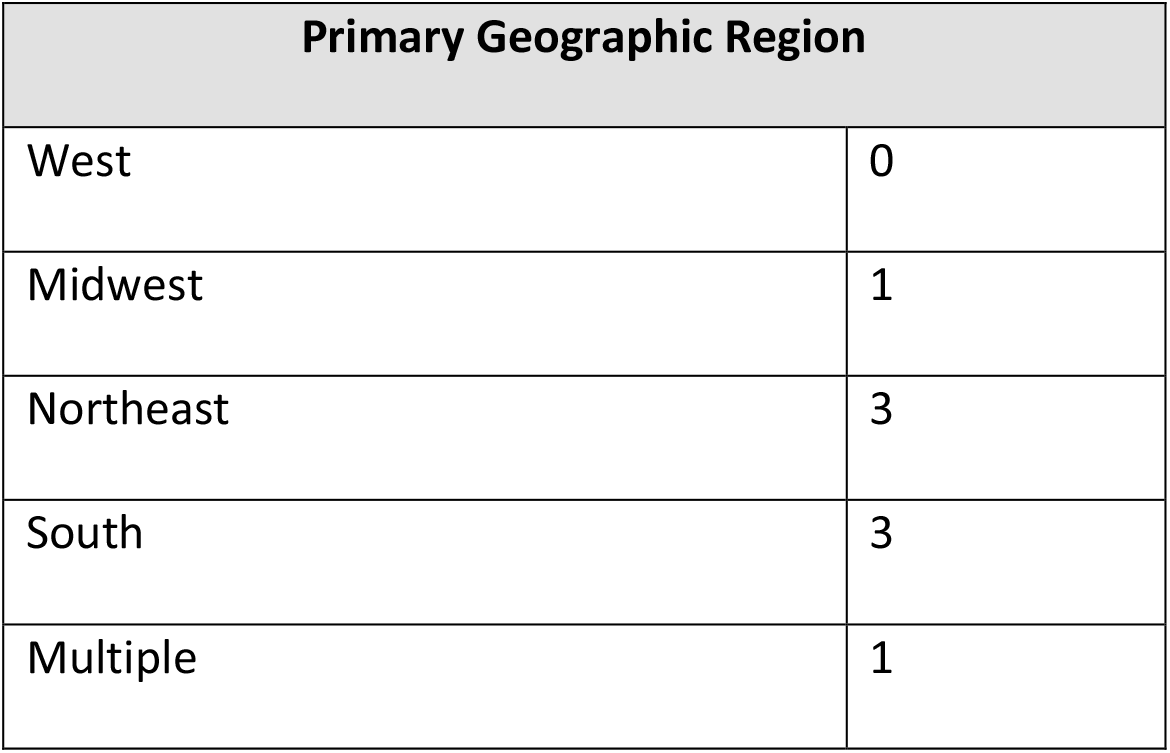
Demographics of Participating Health System (N=8)

| Organizational Structure |  |
| --- | --- |
| Nongovernment, not-for-profit | 8 |
| Number of Hospitals in System |  |
| <5 | 4 |
| 5-10 | 3 |
| >10 | 1 |
| Number of Beds in Hospital System |  |
| <1000 | 1 |
| 1000-2000 | 3 |
| >2000 | 4 |

| Primary Geographic Region |  |
| --- | --- |
| West | 0 |
| Midwest | 1 |
| Northeast | 3 |
| South | 3 |
| Multiple | 1 |

All health systems utilized the same EHR (Epic Systems, Verona, WI). For enterprise resource planning (ERP) systems, four health systems used Infor/Lawson (Infor/ Lawson, New York, NY), one used Oracle/Peoplesoft (PeopleSoft, Pleasanton, CA), and one used SAP (SAP, Germany); data were missing for two health systems.

Three of the eight participating health systems had implemented UDI for implantable devices in one or more designated clinical care areas (e.g., cardiac catheterization labs). One health system was in the process of implementing UDI, while four had implemented UDI in a very limited fashion or not at all.

Characteristics of UDI implementation are shown in **Table 2**. The three health systems that had implemented UDI were using barcode scanning. UDIs were available in EHRs as structured data elements, ERP systems, inventory management systems, and data warehouses. Two additionally had the data available in a database for clinical research. All were capturing UDIs in the cardiac catheterization laboratories. Two were also capturing UDIs in their operating rooms and interventional radiology suites. Point-of-Care information technology systems where UDIs were captured included Pyxis Carefusion, Omnicell Optiflex CL, Epic, and Tecsys.

**Table 2.** Characteristics of UDI Implementation in Health Systems that Have Implemented (N=3)

| Unique Device Identifier Capture Mechanisms |  |
| --- | --- |
| Barcode scanning | 3 |
| Unique Device Identifier Capture Location |  |
| Cardiac catheterization laboratory | 3 |
| Operating room | 2 |
| Interventional radiology | 2 |
| Unique Device Identifier Availability in Information Technology Systems |  |
| Electronic health record | 3 |
| Enterprise resource planning system | 3 |
| Inventory management system | 3 |
| Data warehouse | 3 |
| Patient portal | 2 |

### Qualitative Interviews

Thirty-five individuals completed an interview (20 individual, one dyadic, and four small group). The median interview duration was 50 minutes (range 29, 59). Interviewees represented a variety of position types including seven executives, six directors, four managers, 12 with academic titles, and six individuals who were coordinators and/or support staff. Primary focus areas of interviewees within the health system were supply chain (n=17), clinical research (n=8), informatics (n=4), clinical (n=4), and operations (n=2). Many individuals had more than one focus area and organizational position. Results are reported below in major themes that support or inhibit UDI implementation as well as impact of UDI availability for conducting RWE studies.

Themes 1-4 and associated subthemes are summarized in **Table 3**.

**Table 3:** Themes and Subthemes.

| Theme | Subtheme |
| --- | --- |
| 1. Personal Knowledge of Unique Device Identifier (UDI) |  |
|  | Knowledge of UDI and Current State |
|  | Knowledge of UDI as an Innovation and Attitudes about UDI Priority |
| 2. Organizational Systems, Culture, and Networks |  |
|  | Leadership and Collaboration Between Functional Areas |
|  | Culture of Innovation |
|  | Systems Approach to Technology |
| 3. Factors Outside the Health System |  |
|  | Public Policy Mandates and Supports |
|  | Vendor Technology |
|  | Networks and Organizational Brokers |

| 4. UDI for Generation of Real-World Evidence about Medical Devices |  |
| --- | --- |
|  | Availability of UDIs for RWE Studies |
|  | Lack of UDI Availability for RWE Studies |

### Key Themes

#### Theme 1: Personal Knowledge of UDI

Participants reported that UDI implementation required knowledge among individuals in the health systems, especially individuals with decision authority. This included basic knowledge of what UDI is, the current state of health system implementation, and of UDI as an innovation with potential to improve health system operations.

##### Sub-Theme 1a: Knowledge of UDI and Current State

Among health systems that had implemented UDI, awareness of UDI and its benefits by leadership and key personnel facilitated implementation.

> “*The first set of meetings … was with leadership at [our health system] – … CEOs, CIOs, CMO-level folks to introduce … the concept of UDI*.”

Among health systems that had not implemented UDIs, participants reported a general lack of familiarity with UDI. Operational leaders may assume that since they were tracking devices, their health system had implemented UDI.

Participants in research roles reported additional challenges identifying staff who were knowledgeable about UDI and able to answer questions about what was available in IT systems for RWD analysis.

> “*We have to talk to a lot of different people that have a say about this. We have the clinicians. We have the supply chain people. We have the IT people. We have the business managers. And of all those people, well maybe only the supply chain person really knows what a UDI is*.*”*

##### Sub-Theme 1b: Knowledge of UDI as an Innovation and Attitudes about UDI Priority

Participants noted that leaders and other stakeholders must understand UDI benefits for the health system. That includes operational efficiencies (e.g., supply chain management), regulatory and safety uses (e.g., recall notifications), and comparative effectiveness or safety surveillance research using RWD. Participants from health systems that had implemented UDI shared that portraying how UDI use could improve operational and clinical gaps and inefficiencies surrounding medical devices elicited buy-in.

> *“Recall was primary. three recalls where they had to like go in and manually chart review hundreds and hundreds and hundreds of records and find all the pieces and then contact the people, and so that was a very painful thing*.*”*

Participants from health systems that had not implemented UDI reported difficulty in getting decision makers to see UDI investment as a priority, especially with the financial and other pressures exacerbated by COVID-19.

> “*There’s no business use case right now for UDI adoption. You can’t really show a direct benefit of why this information captured and included in electronic health care records can help you do more or can bring revenue…at this point, anything that we offer to the hospital is about revenue and funding*.”

#### Theme 2: Organizational Systems, Culture, and Networks

The ability of individual proponents and decision makers to implement UDI was described as impacted by systems, both technical and interpersonal, within the organization.

##### Sub-Theme 2a: Leadership and Collaboration Between Functional Areas

Participants described the need to engage stakeholders from various parts of the organization in support of UDI. Some connections opened doors and provided the necessary forum for cross-department collaborations. In some cases, researchers described multiple roles that they held within the organization, and how those varied connections facilitated collaboration with leadership for UDI implementation.

> *“I went to the chiefs of staff and tried to make my case… They were like, ‘Yeah, that’s very valuable. I see the need. We’ve got a list a mile long of things we need to get done, and we’ll sort of try to see what we can do*.*’ But then, what I did next is…coordinate with the Quality and Safety group, and they were much more interested…in safety signals and recalls…Now, they weren’t the primary stakeholder, but they knew all the key stakeholders. And so then, they got me into meeting with supply chain leadership, and I started to talk to them*.*”*

In other health systems, competing organizational priorities and a perceived lack of return on investment for the health system hindered researchers from convincing operational leadership to move forward with UDI implementation.

> *“I spoke to a few senior folks in our organization-those who were dealing with the technology purchase and supply and also, of course, our clinical leaders… just making an argument about scientific perspective is not going to help. Unless you can bring a major grant that will finance it…”*

##### Sub-Theme 2b: Culture of Innovation

Some participants described their health systems as forward thinking and innovative. These organizations prepared for policies and needs that were on the horizon. Organizations saw how the regulation for manufacturers to label devices with UDIs provided an opportunity for them to leverage UDI for improved patient care and billing.

> *“We’ve had very knowledgeable, good leadership across the health system that were following the FDA guidelines [for manufacturers] and the timetable. …already thinking about UDI from the supply and materials management perspective and a technology perspective*.*”*

##### Sub-Theme 2c: Systems Approach to Technology

Organizations that had made a commitment to UDI implementation shared a systems approach towards technology. Some described technology changes (e.g., a new supply chain management system or EHR) as an opportunity to tackle UDI as part of a larger effort. In those cases, research efforts capitalized on the technology change.

> “*The truly digital organizations … didn’t center their world around an EMR and an ERP or a best practice process. What they centered their world around was data management and information and insights, and then they surrounded that with process, and so we had kind of an aha moment…”*

#### Theme 3: Factors Outside the Health System

Participants spoke about the importance of policy mandates, in addition to the value in public-private partnerships and sharing information across health systems. They described facilitators and barriers in relationships outside the health system, including those with vendors, other health systems, public sector partners, and industry associations.

##### Sub-Theme 3a: Public Policy Mandates and Supports

Participants in health systems that had implemented UDIs reported that policy (e.g., 21^st^ Century Cures Act) and the anticipation of future policy (e.g., UDI in claims) played a key role in their health system’s adoption of UDI.

> *“One of [the reasons for implementation] was we understood that the FDA was interested in capturing all that information – that’s #1. And #2, we knew that there were regulatory pressures to make sure that we could track specific implants and items to patients for recalls, any expiration notices, those type of things and so, just in the best interest of our patients, to have a complete record*.*”*

At the same time, participants overall described shortfalls in current policy and the need for stronger policy levers for UDI implementation within health systems and the technology landscape.

> *“The single most important thing would be for CMS to update their billing files so that it was included within it*.*”*
>
> “*It has to be a requirement. It has to be put into the workflow of everybody who’s scanning these devices…”*

Those who had been involved in funded demonstration projects noted the value of those projects in building visibility and momentum within their health system and the opportunity for other health systems if they started using UDI in research.

> “*Making UDI an integral part of every NEST project … might be just enough to get health systems to maybe do UDI just to do a project. And then, when they get into it, they’ll all of a sudden see the benefit and start expanding beyond just the scope of the project, just the way we did*…”

##### Sub-Theme 3b: Vendor Technology

Participants described how advancements in vendor technology were supporting UDI implementation.

> *“When supply chain was ready…to put a new vendor in place, and then there was also an awareness of the utility of this, then it was easy to … make that as part of the vendor selection and contract selection process*.*”*

However, other participants described barriers related to availability of necessary technology and functionality within their health system.

> “*We get the device identifier, but we don’t get the specifics about the manufacturing information … in [our EHR] we do not have a field that is made to hold the UDI as well*.*”*

##### Sub-Theme 3c: Networks and Organizational Brokers

Participants spoke about networking with other organizations or individuals implementing or using UDI in their work.

> *“A group of senior supply chain executives who were meeting were…expressing their frustration about the lack of data standards in the supply chain …they were trying to look for ways of doing things in a much more 21st century sort of way*… *So, that’s what was the impetus …to get these systems together …with the idea of driving standards to the supply chain*.*”*
>
> “*A number of groups … are starting to … center around UDI … it’s got to be captured at the point of care, and they’re trying to figure out how to proceed … NEST is one of those, MDEpiNet, and the [Association for Health Care Resource & Materials Management (AHRMM)] Learning UDI Community*.”

They also spoke about challenges when each organization is tackling UDI implementation and barriers on their own.

> *“There’s a huge amount of waste with each individual hospital and health system maintaining their own Item Master, and the information about the devices*.*”*

#### Theme 4: UDI for Generation of Real-World Evidence about Medical Devices

Participants in health systems that had implemented UDI cited many examples of how UDI availability augmented RWE studies. Those from health systems where UDIs were not available shared limitations in performance of RWE studies. (**See Table 4 for representative quotes**).

**Table 4:**
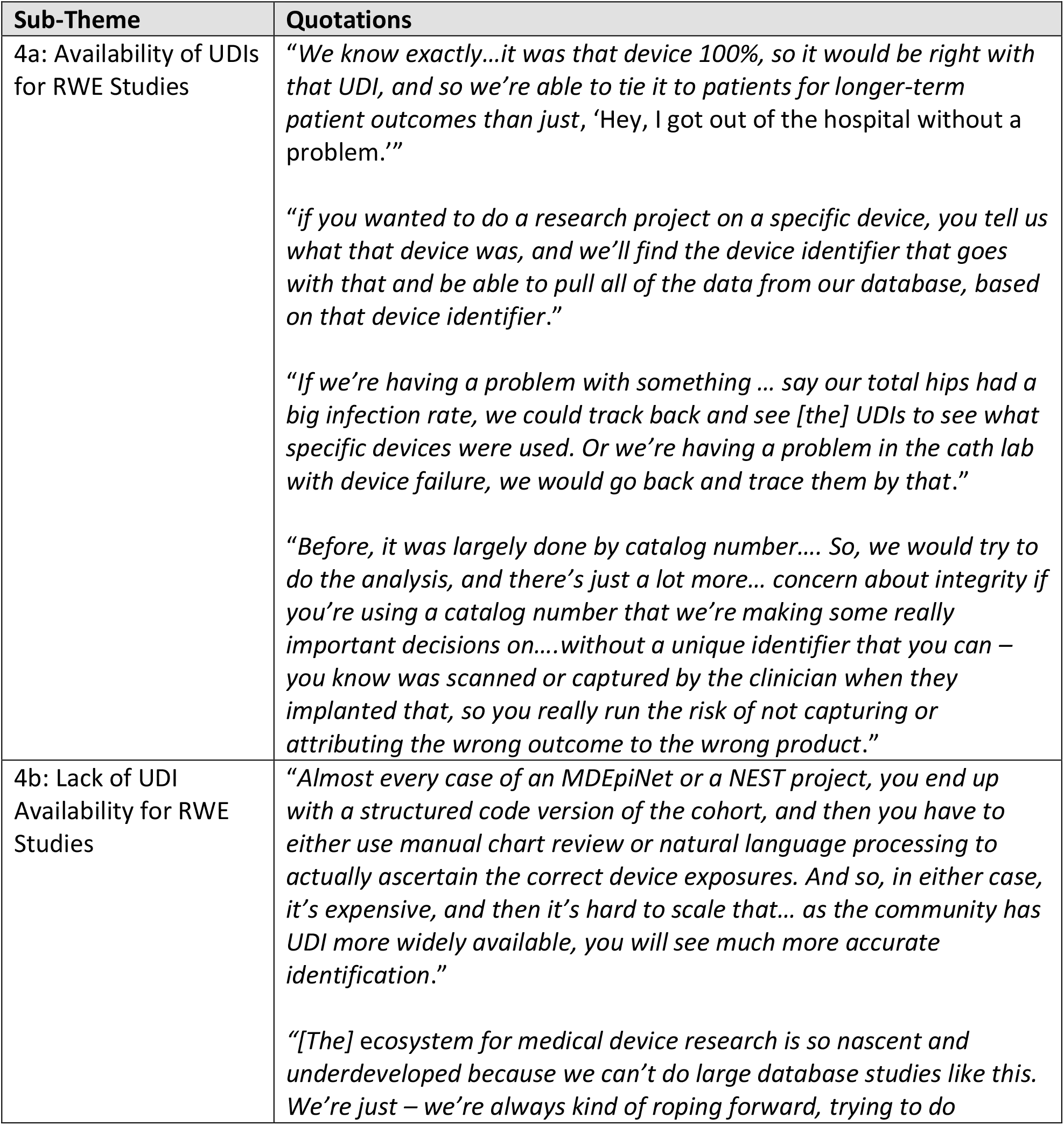
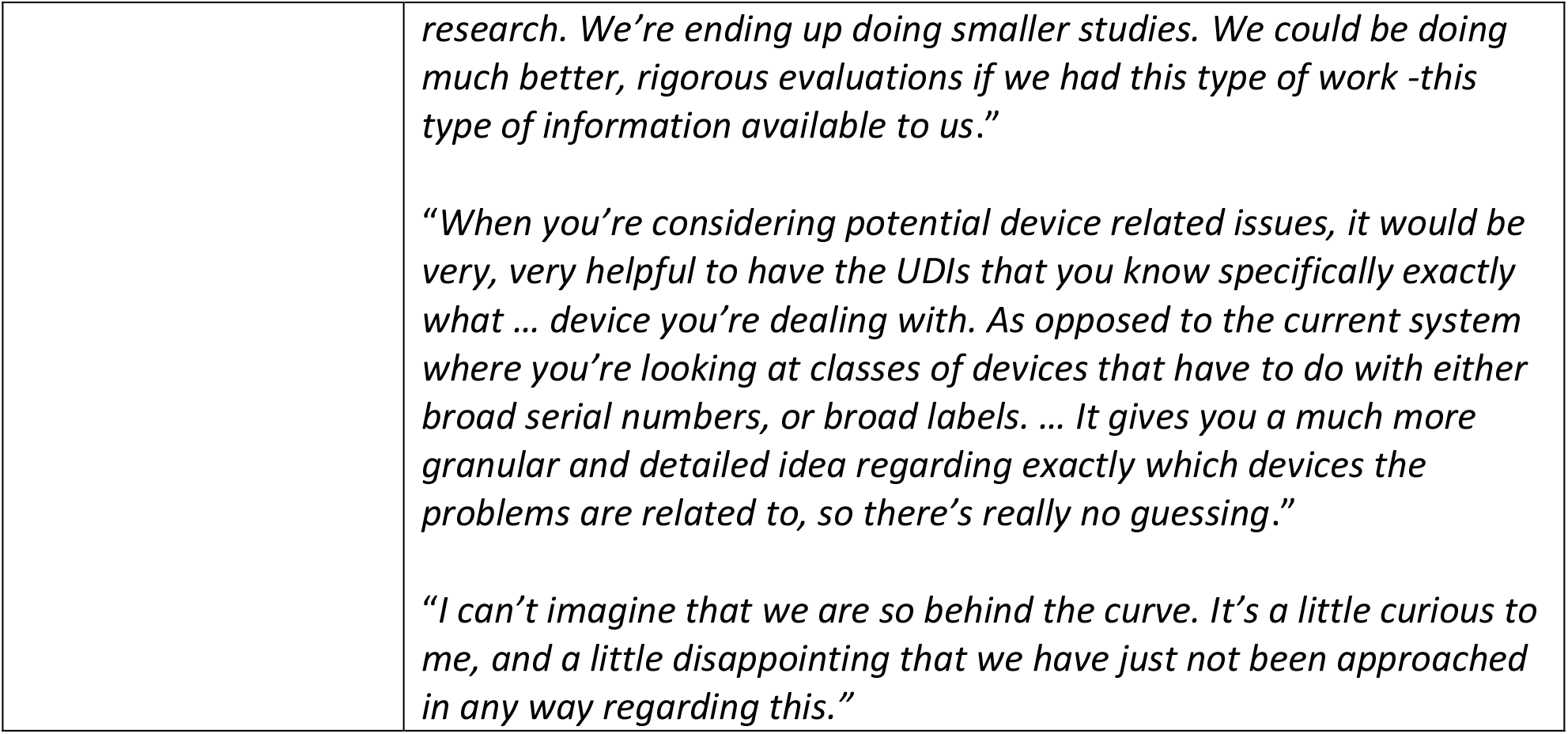
Theme 4-UDI for Generation of Real-World Evidence about Medical Devices with Associated Sub-themes and Exemplar Quotes.

| Sub-Theme | Quotations |
| --- | --- |
| 4a: Availability of UDIs for RWE Studies | *“We know exactly...it was that device 100%, so it would be right with that UDI, and so we’re able to tie it to patients for longer-term patient outcomes than just, ‘Hey, I got out of the hospital without a problem.’”*<br>*“if you wanted to do a research project on a specific device, you tell us what that device was, and we’ll find the device identifier that goes with that and be able to pull all of the data from our database, based on that device identifier.”*<br>*“If we’re having a problem with something ... say our total hips had a big infection rate, we could track back and see [the] UDIs to see what specific devices were used. Or we’re having a problem in the cath lab with device failure, we would go back and trace them by that.”*<br>*“Before, it was largely done by catalog number.... So, we would try to do the analysis, and there’s just a lot more... concern about integrity if you’re using a catalog number that we’re making some really important decisions on....without a unique identifier that you can – you know was scanned or captured by the clinician when they implanted that, so you really run the risk of not capturing or attributing the wrong outcome to the wrong product.”* |
| 4b: Lack of UDI Availability for RWE Studies | *“Almost every case of an MDEpiNet or a NEST project, you end up with a structured code version of the cohort, and then you have to either use manual chart review or natural language processing to actually ascertain the correct device exposures. And so, in either case, it’s expensive, and then it’s hard to scale that... as the community has UDI more widely available, you will see much more accurate identification.”*<br>*“[The] ecosystem for medical device research is so nascent and underdeveloped because we can’t do large database studies like this. We’re just – we’re always kind of roping forward, trying to do* |
|  | *research. We're ending up doing smaller studies. We could be doing much better, rigorous evaluations if we had this type of work -this type of information available to us."*<br>*"When you're considering potential device related issues, it would be very, very helpful to have the UDIs that you know specifically exactly what ... device you're dealing with. As opposed to the current system where you're looking at classes of devices that have to do with either broad serial numbers, or broad labels. ... It gives you a much more granular and detailed idea regarding exactly which devices the problems are related to, so there's really no guessing."*<br>*"I can't imagine that we are so behind the curve. It's a little curious to me, and a little disappointing that we have just not been approached in any way regarding this."* |

##### Sub-Theme 4a: Availability of UDIs for RWE Studies

Researchers from health systems that had implemented UDI within their data systems discussed the benefits for generation of RWE on medical devices. Devices could be accurately identified, linked to patients, and clinical outcomes could be studied.

##### Sub-Theme 4b: Lack of UDI Availability for RWE Studies

Researchers at health systems without UDI availability discussed the challenges faced, including cumbersome manual processes to identify devices and outcomes. These researchers voiced a desire for UDI implementation, which would allow for larger, more robust studies using RWD.

## DISCUSSION

In this mixed methods study of UDI implementation in NESTcc health system network collaborators, we found heterogeneity in UDI implementation and its use in RWE studies. Three participating health systems were mature in UDI implementation. Whereas clinical and operational benefits - not RWE generation - catalyzed UDI implementation in these health systems, RWD with UDIs was available for use in medical device safety and effectiveness evaluations. Participants shared the benefits for research including greater data integrity, linkage to patient-level data, and efficiency in use of device-specific data. Half of the participating health systems had not implemented UDI or had done so in a very limited fashion. Researchers in these systems reported that conducting RWE-based research was cumbersome, costly, and not scalable because of challenges in medical device identification. To our knowledge, these findings provide the first juxtaposition of experiences of researchers who have and do not have UDIs available for RWE studies.

In the health systems that had adopted UDIs, implementation was facilitated by organizational UDI knowledge, interdisciplinary collaborations, and a systems approach to innovation and technology. Leaders and staff in these health systems were engaged in external UDI education and collaborative opportunities. They stayed current on relevant policy and were persistent in their navigation of external factors, particularly the IT functionality needed for their UDI implementation and use. External collaborations such as the AHRMM Learning UDI Community and the Strategic Marketplace Initiative (SMI) were very valuable for knowledge, collaboration, and support. Researchers involved in medical device evaluations in these health systems collaborated with FDA as well as colleagues in other health systems and served as organizational brokers for UDI implementation efforts, thus supporting the feasibility of UDI availability for RWE studies.

In contrast, in the other studied health systems, UDI adoption was hindered by a lack of organizational knowledge of UDI and its benefits, barriers to building organizational collaboration surrounding UDI, and lack of support for the initial investment. Lack of external policy requiring UDI implementation imposed an immediate barrier. Researchers interested in medical device evaluations in these health systems were frustrated by the lack of UDI availability.

Our findings on the limited availability of UDIs in health systems for RWE are consistent with the literature in this nascent area^15 16^ as are our findings on overall UDI implementation,^19 21-23^ particularly the fundamental importance of awareness and education on UDI; knowledge of UDI’s purpose and innovation; the role of leaders, expertise, and relationships across functional units in a health system; and relationships external to the health system. IT and policy barriers have been an important focus of discussion in the literature and professional forums.^19 21 23 31^ Our findings on the benefits of UDI use in RWE studies coupled with published literature where UDIs have been used in RWE studies^15 32-34^ put a spotlight on the centrality of UDI for accurate device identification and linkage of RWD sources.

Ultimately, realizing the public health goals of the UDI Final Rule and goals to use RWD to evaluate medical device safety and effectiveness requires widespread adoption of the UDI in health systems.^23^ Despite significant regulatory efforts and advancements through the UDI Rule and creation of NESTcc, a critical step for robust RWE studies on medical devices is lacking – adoption of UDIs in health systems. More health systems with UDI availability need to participate in order to derive the benefits of distributed research that NESTcc has been set up to support.^9^

There are some important limitations of our study that deserve consideration. We were unable to obtain interviews with some health systems, which may limit generalizability. Some health systems declined participation indicating they had not implemented UDIs, so felt they had nothing to share on the topic. Others indicated they were too busy to participate. However, a majority of eligible health systems participated. We also were unable to interview all identified potential individual participants. Some did not feel adequately informed about UDI implementation to provide comprehensive information about their health system. This was addressed in some cases by conducting dyadic, triatic, or quadratic interviews in which multiple people from the health system provided their perspective. Additionally, the first point of contact, the NESTcc research leads, did not always have connections with personnel in supply chain management or IT. It is possible that individuals with expertise in UDI within health systems were not identified, despite chain-referral sampling. However, our experience likely represents a best-case scenario of knowledge and understanding of UDI adoption in health systems to support RWE.

In conclusion, our findings highlight that UDI implementation within health systems needs to be supported and advanced to achieve the goals for RWE generation on medical devices. NESTcc is well-poised to facilitate the effort to advance UDI adoption in other NESTcc health system network collaborators by capitalizing on the expertise of those health system network collaborators mature in UDI implementation and UDI availability for RWE studies, along with advancing a more generalizable playbook for UDI implementation in health systems.

## Supporting information

SRQR Checklist

## Data Availability

Data are not available given confidentiality provisions as part of informed consent.

## Acknowledgements

This project was supported by a research grant from the Medical Device Innovation Consortium (MDIC) as part of the National Evaluation System for health Technology (NEST), an initiative funded by the U.S. Food and Drug Administration (FDA) through grant 1U01FD006292-01. Its contents are solely the responsibility of the authors and do not necessarily represent the official views nor the endorsements of the Department of Health and Human Services or the FDA. While MDIC provided feedback on project conception and design, the organization played no role in collection, management, analysis and interpretation of the data The research team, not the funder, made the decision to submit the manuscript for publication.

Views expressed in written materials or publications and by speakers and moderators do not necessarily reflect the official policies of the Department of Health and Human Services; nor does any mention of trade names, commercial practices, or organization imply endorsement by the United States Government.

## Author Contributorship

All named authors meet the International Committee of Medical Journal Editors (ICMJE) criteria for authorship for this article. Manuscript writing: SSD and NAW. Manuscript revision for important intellectual content: JLR, JSR, JPD.

