## Supplementary Material 1 for "Exploring Unique Device Identifier Implementation and Use for Real-World Evidence: A Mixed-Methods Study with NESTcc Health System Network Collaborators"

**Pre-Interview Survey**

These survey questions are aimed at hospitals, or hospital systems, about their adoption and use of the unique device identifier (FDA UDI). Please answer this survey on behalf of specific hospitals or clinical settings that have the greatest adoption of the UDI. If you are discussing a specific hospital(s) please indicate that in the Comments section.

**Hospital System: ___________________**

**A. General**

**Organizational Structure**

- o Federal government
- o Non-federal government
- o Not-for-profit, non-government
- o For-profit, investor-owner

**Are there multiple hospitals as part of the health system?**

- o Yes
    - If yes, how many hospitals? __
- o No

**Number of licensed beds across health system**: __

**Number of patients cared for in 2020**: __

**B. Information Technology Systems**

**What is the EHR vendor(s)? Check all that apply**

- o Epic
- o Cerner
- o Other: __________

**What is the Enterprise Resource Planning (ERP) vendor?**

- o Infor/Lawson
- o Oracle/Peoplesoft
- o Workday
- o SAP
- o Other: ___________

**Are you using any Point of Care (POC) or Point of Use (POU) system to scan device, supply, or implant capture?**

**If yes, continue to Section C.**
**If no, skip to Section F.**

**C. UDI Capture**
**What is the primary method of capture of the UDI? Check all that apply:**

- o Barcode scanning
- o RFID
- o Manual entry (keyboard)
- o Other_____________________________
- o Not capturing

**At any of the hospitals within your hospital system, are UDIs for implantable devices captured and documented at any of the following procedural sites? (Please answer all that are applicable and "N/A" if unaware).**

| **Procedural Site** | **UDI Captured and Documented (Yes/No)** | **If Yes, approximate percentage of sites (1-100%)** | **Point-of-care system with UDI** |
|---|---|---|---|
| **Cardiac catheterization laboratory** | | | |
| **Operating Room** | | | |
| **Interventional Radiology** | | | |
| **Other (please name)** | | | |
| **Other (please name)** | | | |
| **Other (please name)** | | | |
| **Other (please name)** | | | |

**D. UDI Stored (either PI or DI)**
**Indicate UDI availability in any of the following. Check all that apply.**

- o Electronic health record(s)
- o ERP system
- o Inventory management system(s)
- o Data warehouse
- o Database for clinical research or QI projects (e.g. NESTcc test cases)
- o Clinical registries. Please indicate which __________________
- o Patient portal
- o Other _________________________

**If the UDI is available in the electronic health record(s), is it available as a structured data element (i.e., a data element that can be easily and accurately abstracted and coded identically across records)?**

- Yes
- No

**If relevant, indicate from which IT system (e.g. EHR, point-of-care system) the UDI is transferred to the following data repositories?**

- Data warehouse ______________________
- Database used for device-related research_____________
- Clinical registries (e.g. American Joint Replacement Registry/AJRR) ___________________

**E. Comments:**

_____________________________________________________________________________

_____________________________________________________________________________

_____________________________________________________________________________

**F. Referral of Interview Participants**

We would like to interview individuals about UDI implementation, adoption, and use in your hospital system particularly focused on clinical procedural areas.

**Please provide the name and contact email of at least four individuals in the areas below, who have a significant role in UDI implementation. Please feel free to send the survey back even if you do not have names of individuals yet; you can send us the names below to XX email address whenever you have them.** Even if UDI is not yet implemented or is implemented in a limited way in your organization, we are still very interested in talking with individuals in your organization who are knowledgeable of medical device documentation, identification, tracking and use of this data.

- Supply Chain Management - such as a Physician or other Director
- Information Technology – such as the Chief Medical Information Officer, IT manager
- High-volume procedural areas (cath lab, OR, IR) – such as administrative or clinical leader
- Formal or informal leader or champion supporting UDI implementation efforts
- Others – such as Director of quality improvement, director of recalls, or individuals involved in risk management

| Name | Title | Email Address |
|---|---|---|
