## Supplementary Material 2 for "Exploring Unique Device Identifier Implementation and Use for Real-World Evidence: A Mixed-Methods Study with NESTcc Health System Network Collaborators"

### Evaluation of Uptake of Unique Device Identifiers (UDIs) by Health Systems

#### Interview Guide

**Section 1: Introduction**

1) The goal of this interview is to understand UDI implementation and use in clinical care in your health system. So that I can better understand the perspective that you're sharing with us, can we start by having you tell me what your positions/roles is within the health system *(Probes: clinical, IT, supply chain, administration, other),* as well as how your role/department is involved in UDI implementation, adoption and use?

**Section 2: UDI availability and implementation**

2) How would you describe the state of UDI implementation in your health system at this point in time? if so, how? [If none → skip to Q6]
   - *Probe: Which systems (e.g., supply chain* ("ERP/MMIS")*, EHR, and/or other specific information systems such as cardiac catheterization lab or EP lab software; entire UDI or just the DI or PI)*
   - *Probe: Which clinical areas or sites/hospitals?*
   - *Probe: Which devices (e.g., implantable, non-implantable)?*
3) What were some of the reasons that UDI implementation efforts started in these areas? *(Probes: who/what was behind the first initiative, , was it for implantable or non-implantable devices, what was the 1st clinical site, how/why did it advance beyond the 1st site)*
   a) How would you describe the reason behind UDI implementation (in that site[s])? How would you describe the purpose it hoped to achieve or the problem it was meant to address?
4) We are trying to understand who or what is needed for health systems to be successful at implementing UDI (or what—if missing—would make UDI implementation harder). What types of support or infrastructure facilitated your organization's UDI efforts? *(Probes: financial, organizational, IT-related, clinical, facilities?)*
   a) Was there a particular catalyst—a person or an event—that got you started or pushed your progress even further? [If yes], can you tell me about that and why you think it made the difference?
   b) Can you tell me how the case was made for UDI implementation among leadership? (*Probe: specific trigger(s)*)
   c) Can you tell me how staff buy-in was approached? What was effective? *(Probes: workflow alteration, training)*
   d) Can you tell me how the IT needs were addressed for your health system? (*Probes: creation of a device database, modification of an existing database, interoperability*)
      - What sources of data are you using to support it?
   e) Can you tell me how the necessary IT infrastructure was prioritized? (*Probe: Is the infrastructure complete, including interoperability?*)
5) What challenges were faced in UDI implementation at clinical sites, and how were they worked through? *(Probes: financial, organizational, IT-related, clinical, facilities?)*

**Note: Question 6 is intended to address the situation if the UDI is completely unavailable (i.e., "no" answered to question 2)**

6) Do you know why UDIs are not available in your organization at clinical workflows? (*Probe:* Were there specific obstacles that were faced?)
    a) Has there been interest in UDI implementation & use? If so, for what purpose?

**Section 3: Current medical device tracking & documentation**

7) How are medical devices currently tracked within the clinical supply chain?
    a) Are there differences between implantable and non-implantable devices (e.g., catheters)?
    b) Are there other distinctions across devices that characterize the process of device tracking?
    c) If the UDI has been used, how has it been helpful? How does the process with UDI compare to the process without UDI?
    d) Are there processes or challenges that could be improved with UDI integration (or greater UDI integration)?
8) How do you ensure that expired or recalled devices are not used in clinical care? That devices are re-ordered when supply is needed?
    a) If the UDI has been used, how has it been helpful? How does the process with UDI compare to the process without UDI?
9) How is medical device information currently documented in procedure reports and other clinical notes?
    a) Are there differences between implantable and non-implantable devices?
    b) Are there other distinctions across devices that explain device documentation?
    c) If the UDI has been used, how has it been helpful? How does the process with UDI compare to the process without UDI?
    d) What are the challenges in documenting medical device information?
    e) Are there processes or challenges that could be improved with UDI integration (or greater UDI integration)?
10) How did having or not having the UDI impact tracking of devices or equipment during the disruptions related to COVID-19?
    a) What was the value of the UDI?
    b) If not, how could tracking be better addressed?
    c) Was there other value and uses of UDI during the COVID-19 pandemic?

**Section 4: Desire and vision for UDI implementation**

11) Are there clinical areas where you want to implement UDI but haven't been able to yet? If so, can you tell me about it?
    a) What happened and what could be changed in order to successfully implement there?
    b) What do you think are reasons it hasn't gotten traction in these areas?
12) Are there future plans for any additional UDI implementation?
    a) What would be necessary to support greater adoption in your health system? (*Probes: Policies, e.g. integration into CMS claims and requirement of the DI for billing? Hospital system goals for supply chain modernization? Evidence, e.g.,*

*Improved data analysis and assessment of device-requiring procedures? Return-on-investment analysis? Education, e.g. greater awareness among key decision-makers?)*

b) What values or standard of performance need to be recognized to make this a priority?

c) What support could be provided, and by whom? *(Probes: by hospital system, C-suite in health system leadership [CMO, CNO, COO, CFO], manufacturers, EHRs, ERP vendors, labelers, champion)*

13) What is needed to move UDI adoption forward in the U.S.?

a) Where are the biggest challenges?

b) What are the greatest opportunities?

**Section 5: UDI Use**

14) We would now like to discuss various uses of the UDI. Please answer only those questions about which you have information about uses.

a) First, we would like to discuss recalls. If you have been involved in recalls, we ask you to consider the most recent recall(s)

- Is UDI being used?
  - a) If yes, how does its use benefit the recall process?
  - b) Do you have documented benefit of use (e.g., error reduction, time savings, cost savings, etc.)?
- If not, how was the device identified?
- How would availability of the UDI better support a medical device recall?

b) We would now like to discuss use in supply chain management?

- Is UDI being used?
  - a) If yes, how does it benefit the supply chain team projects?
  - b) Do you have documented benefit of use? (e.g., inventory optimization? Time saved?

c) We would now like to discuss research or quality improvement projects focused on a specific medical device (e.g., NESTcc Test-Case)

- Is UDI being used?
  - c) If yes, how does it benefit the projects?
  - d) Do you have documented benefit of use? (e.g., aggregation of device data, larger datasets, etc.)
  - e) Has the UDI been used outside of NESTcc test-cases?
- If not, how have medical devices been identified?
- How would availability of the UDI better support your QI or research studies of medical devices?

d) We would now like to discuss clinical care and patient knowledge about their device(s).

- How is UDI being used from a patient perspective? (*Probes: in patient portal, on implant card, in transfer of health information, etc*.)
- How is UDI being used from a clinician perspective? (*Probes: available in EHR for use prior to revision procedures, on implant card, in transfer of health information, etc*.)
- If used, how does it benefit clinical care? Do you have documented benefit of use?

- If not used, how would availability of UDI better support patient care?

e) We would now like to discuss clinical registries.
- Is UDI being used? If so, what have been the benefits?
- Are there challenges to including UDI?
- How would use of UDI better support registry data?

f) Are you aware of any other uses of UDI?
- Do you have documented benefit of use?

**Section 6: Metrics and Other Benefits**

15) What financial or clinical metrics could be better served with availability and use of UDI for your health system? *(Probes: cost of expired devices, recalled devices, devices that malfunctioned during deployment, reporting to MAUDE, any potential quality metrics, academic manuscripts/publications)*
16) What other benefits would you anticipate for your health system with UDI use?
    a) What metrics will be monitored by decision-makers? (e.g., product standardization, reduced inventory on hand and increased utilization of devices that are purchased for cost savings, other financial metrics, safety metrics, other?)

**Section 7: Closing**

17) Is there anything else you think we should know?
18) Is there anyone else with whom you think we should be talking with about these topics?
